# Capturing intrahost recombination of SARS-CoV-2 during superinfection with Alpha and Epsilon variants in New York City

**DOI:** 10.1101/2022.01.18.22269300

**Authors:** Joel O. Wertheim, Jade C. Wang, Mindy Leelawong, Darren P. Martin, Jennifer L. Havens, Moinuddin A. Chowdhury, Jonathan Pekar, Helly Amin, Anthony Arroyo, Gordon A. Awandare, Hoi Yan Chow, Edimarlyn Gonzalez, Elizabeth Luoma, Collins M. Morang’a, Anton Nekrutenko, Stephen D. Shank, Peter K. Quashie, Jennifer L. Rakeman, Victoria Ruiz, Lucia V. Torian, Tetyana I. Vasylyeva, Sergei L. Kosakovsky Pond, Scott Hughes

## Abstract

Recombination is an evolutionary process by which many pathogens generate diversity and acquire novel functions. Although a common occurrence during coronavirus replication, recombination can only be detected when two genetically distinct viruses contemporaneously infect the same host. Here, we identify an instance of SARS-CoV-2 superinfection, whereby an individual was simultaneously infected with two distinct viral variants: Alpha (B.1.1.7) and Epsilon (B.1.429). This superinfection was first noted when an Alpha genome sequence failed to exhibit the classic S gene target failure behavior used to track this variant. Full genome sequencing from four independent extracts revealed that Alpha variant alleles comprised between 70-80% of the genomes, whereas the Epsilon variant alleles comprised between 20-30% of the sample. Further investigation revealed the presence of numerous recombinant haplotypes spanning the genome, specifically in the spike, nucleocapsid, and ORF 8 coding regions. These findings support the potential for recombination to reshape SARS-CoV-2 genetic diversity.

## INTRODUCTION

Recombination is a common evolutionary feature of positive-strand RNA viruses^1^. It can increase genetic diversity and accelerate adaptation in viral populations by combining existing linked allelic variation. The signature of frequent recombination is pervasive across Betacoronaviruses in bats and other animal ^2-6^ hosts, and its detection is made easier in part by the substantial genetic divergence separating these various coronaviruses. When an individual is simultaneously infected with two genetically distinct strains of a virus, so-called superinfection ^7^, these viruses can recombine to produce a virus with novel allelic combinations. Although recombination is expected to regularly occur during SARS-CoV-2 infections, it can be difficult to detect *in vivo* unless it involves genetically distinguishable parental strains: recombination between two identical or nearly identical genomes leaves no detectable molecular trace. Furthermore, as with influenza virus ^8,9^, SARS-CoV-2 superinfections have been only rarely reported in humans ^10-12^, likely due to the short mean duration of SARS-CoV-2 infections.

Early claims of recombination in SARS-CoV-2 ^13^ may have been confounded by sequencing errors or convergent evolution ^14^. As the COVID-19 pandemic progressed, and genetically divergent lineages have evolved, evidence for recombination between these lineages is becoming more convincing ^15,16^. Many of these divergent lineages include highly transmissible variants distinguished by S genes that, under positive selection, have accumulated multiple mutations associated with increased transmissibility, virulence, and immune escape ^17,18^.

Here, we provide a detailed characterization of an instance of superinfection from January 2021, identified by the New York City (NYC) Department of Health and Mental Hygiene (DOHMH). We show that this individual was superinfected with two SARS-CoV-2 variants: Alpha (B.1.1.7) and Epsilon (B.1.429) ^19,20^. Further, we characterize evidence for recombination occurring within this superinfected individual, providing an *in vivo* snapshot of this evolutionary process within SARS-CoV-2.

## RESULTS

### Patient epidemiology

In December 2020, researchers and public health officials in the United Kingdom identified a rapidly spreading SARS-CoV-2 variant within England, then designated as PANGO lineage B.1.1.7 ^20^, now designated as the Alpha variant of concern in the WHO nomenclature. In NYC, a SARS-CoV-2 genome sequence classified as belonging to the Alpha lineage was obtained from a sample drawn on 4 January 2021 (the ‘index case’). Due to the potential public health importance of Alpha variant cases in NYC in early 2021, NYC DOHMH conducted a public health investigation related to the individual from which this sample had been obtained: NYCPHL-002130 (GISAID accession number *EPI_ISL_857200*). This investigation determined that the individual had recently traveled to Ghana (late December/early January), and contact tracing identified another case of an Alpha variant infection, sampled on 14 January 2021, in a named contact with a similar travel history (the ‘named contact partner’): NYCPHL-002461 (GISAID accession *EPI_ISL_883324*).

### Atypical Alpha (B.1.1.7) variant PCR screening

Typical of the Alpha variant ^20^, NYCPHL-002130 exhibited S gene target failure (SGTF) phenotype with the TaqPath COVID-19 RT-PCR assay (Table 1). NYC PHL uses the ARTIC amplicon-based protocol V3 to sequence full viral genomes and capture intrahost diversity. The viral genome from this index case showed limited intrahost viral diversity (Figure 1). A single variable site was found at position 23099, with C in 20.4% of reads and A in 79.6% of reads. All other substitutions differentiating this sequence from the Wuhan-Hu-1 reference genome sequence (*NC_045512*.*2*) were present in >99.0% of reads.

**Table 1.** Cycle threshold (Ct) values from TaqPath assays from index case (NYCPHL-002130) and named contact partner (NYCPHL-002461).

| Case | WGS ID | Ct value |  |  |
| --- | --- | --- | --- | --- |
|  |  | ORF1ab | N gene | S gene |
| Index case | NYCPHL-002130 | 14.97 | 15.44 | N/A |
| Named contact partner | NYCPHL-002461-A | 14.86 | 15.70 | 17.34 |
|  | NYCPHL-002461-B | 16.06 | 16.251 | 18.68 |
|  | NYCPHL-002461-C | N/A | N/A | N/A |
|  | NYCPHL-002461-D | 15.73 | 15.83 | 18.35 |

**Figure 1.**
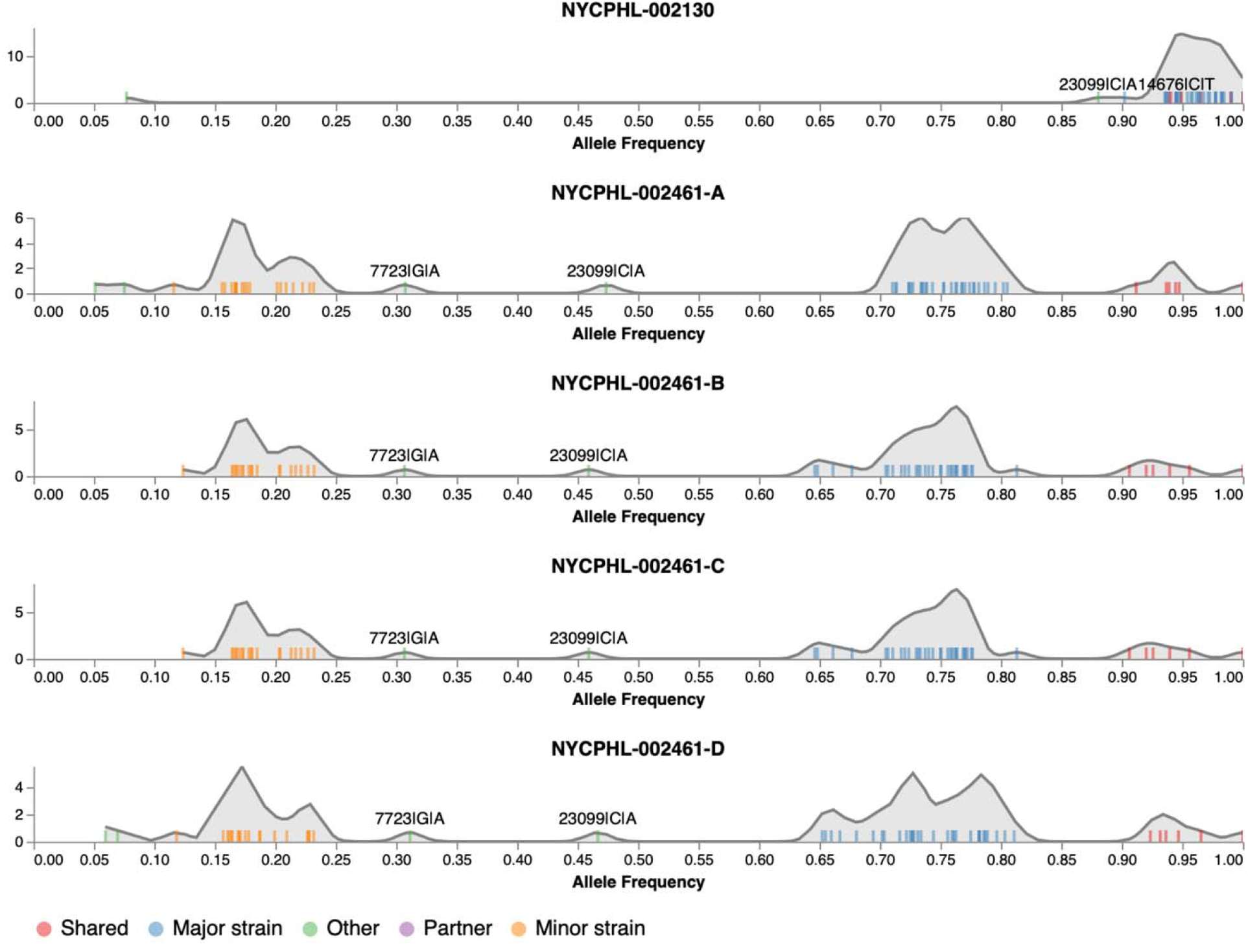
The distribution of allelic frequencies in the index case (NYCPHL-002130) and named partner with suspected superinfection (NYCPHL-002461). Frequencies of individual alleles shown as ticks, a smoothed kernel density plot is used to highlight clustering patterns, and colors represent allele types.

During the initial PCR screening of the sample collected from the named contact partner (NYCPHL-002461), the SGTF characteristic of the Alpha variant was not observed (Table 1). Furthermore, genome sequencing revealed substantial intrahost viral diversity within the viral genome, a possible signature of superinfection (Figure 1). To confirm that this intrahost diversity was not attributable to experimental or sequencing artifacts, the original sample was re-extracted and re-sequenced (NYCPHL-002461-B) and similar SGTF was observed. Additional extractions were then performed in duplicate from the original stock (NYCPHL-002461-C and - D) and sequenced. The same signature of intrahost diversity was confirmed in all four sequenced extractions. Four nucleotide (nt) substitutions differentiating this sequence from the reference genome were identified at >90% frequency: C241T, C3037T, C14408T, and A23403G. Numerous additional substitutions, including A23063T (S N501Y), were present, but at slightly lower frequencies. Nonetheless, this genome was classified as an Alpha variant. Notably, the Δ69/70 and Δ144 deletions were found at >95% in the sequencing reads, despite the lack of SGTF.

NYCPHL-002461-A, -B, and -D extracts exhibited low Ct values for the ORF1ab and N gene targets, ranging between 15 and 16 (Table 1). The S gene target Ct values were around 2 to 3 cycles higher. The difference suggests a reduction of viral template in the S gene target region, but not SGTF. We note NYCPHL-002461-C yielded an invalid result, as the TaqPath assay showed no amplification on all targets, including the MS2 phage extraction-control target.

### Intrahost diversity

The presence of multiple intermediate frequency alleles and the lack of SGTF in the TaqPath assay prompted us to investigate the intrahost diversity in the named contact partner, NYCPHL-002461. Major and minor intrahost strains were distinguished using the previously described and validated Galaxy SARS-CoV-2 allelic variation pipeline ^21^.

The four replicate sequencing runs for NYCPHL-002461 yielded remarkably similar patterns with allelic frequencies segregating into four categories: shared, major strain, minor strain and other (see Figure 1, interactive notebook at https://observablehq.com/@spond/nyc-superinfection). Shared alleles were those present at 90% allele frequency (AF) in three or more samples. The four shared alleles constituting substitution mutations are the same substitutions that define the ancestor of the Lineage B viruses circulating in the United States: C241T, C3037T, C14408T, A23403G (Figure 1; Supplementary Table 1). Two deletions in the S gene (Δ69-70 and Δ144) were also present at high AF (>97%).

Major strain alleles are those that occurred at frequencies between 60 and 90% (≥3 samples), with all diagnostic Alpha mutations in this set. Minor strain alleles are those that occurred at frequencies between 10 and 25% (≥3 samples), with all but one diagnostic Epsilon mutation in this set; the A28272T mutation characteristic of Epsilon is absent in NYCPHL-002461. Notably, the “other” category encompasses all other variable sites, i.e. those occurring at AF between 25% and 60% or those found in only one or two samples. The two alleles were found in all four replicate sequences at intermediate frequencies: G7723A (30.3%) and C23099A (46.7%). These frequencies are suggestive of intrahost variation in the major strain.

In contrast, the sequencing dataset for the index case, NYCPHL-002130, showed all but one of the alleles occurring at ≥85%, and all but one of the alleles (C14676T) were also found as “shared” or “major strain” classes in the NYCPHL-002461 datasets (Figure 1). The C23099A mutation, which was at intermediate frequency in NYCPHL-002461, was present at only 88.1% in NYCPHL-002130 from the index case, suggesting the transmission of a mixed viral population between these individuals.

### Phylogenetic inference with major and minor variants

We identified sub-clades within Alpha and Epsilon that shared substitutions with the major and minor strains (Figure 2). We inferred a maximum likelihood (ML) phylogenetic tree in IQTree2 for the major strain and 1174 related B.1.1.7 genomes containing the C2110T, C14120T, C19390T, and T7984C substitutions found in the major strain (Figure 2A). We also inferred an ML tree for the minor strain and 807 related B.1.429 genomes containing the C8947T, C12100T, and C10641T substitutions found in the minor strain (Figure 2C).

**Figure 2.**
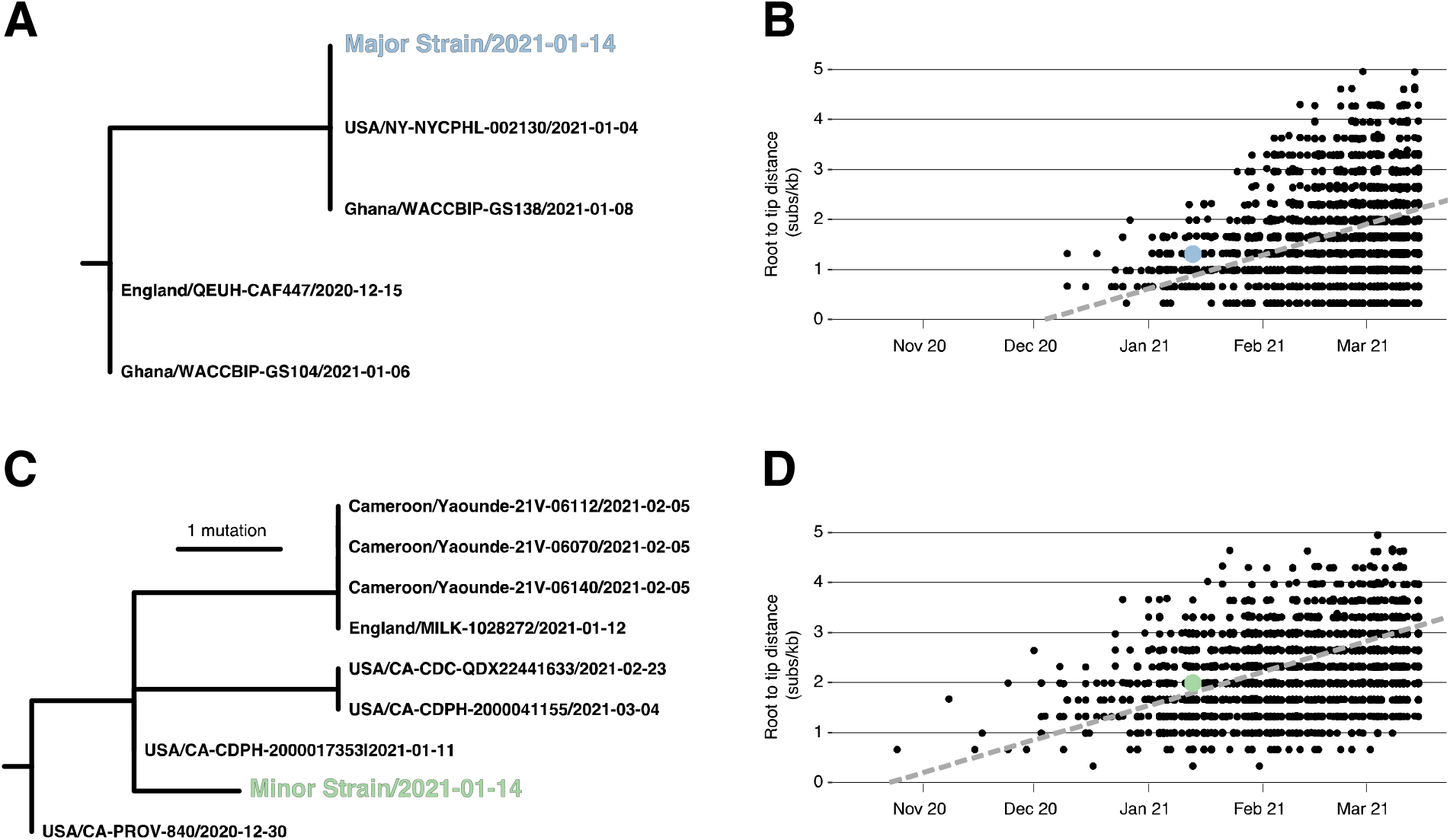
Phylogenetic consistency of major and minor variants. (A) Phylogeny of B.1.1.7 immediate relatives, (B) Root-to-tip regression for B.1.1.7, (C) Phylogeny of B.1.429 immediate relatives, (D) Root-to-tip regression for B.1.429. NY-NYCPHL-0024661 is the genome deposited in GISAID from the case of putative superinfection.

Root-to-tip regression analyses show that the NYCPHL-002461 sampling date is consistent with the molecular clock for both the major and minor strain sequences (Figure 2B/2D), indicating that one would expect viruses of this degree of genetic divergence to have been circulating in mid-January 2021. In fact, genomes identical to the major variant were sampled in both NYC (the NYCPHL-002130 index case) and in Ghana (EPI_ISL_944711) on 8 January 2021, consistent with a scenario in which this particular Alpha virus was introduced into NYC by an individual who had recently traveled to Ghana and had contact with the named contact partner. These three viruses share a common ancestor around 4 January 2021 and are separated by additional viruses sampled in Ghana by two mutations: C912T and C23099A. Notably, the latter mutation appears at intermediate frequency in both NYCPHL-002130 and NYCPHL-002461.

The minor variant is genetically distinct from all other sampled genomes, including any genome sequenced by NYC DOHMH (Figure 2C). The closest relatives were sampled in California (EPI_ISL_3316023, EPI_ILS_1254173, EPI_ISL_2825578), the United Kingdom (EPI_ILS_873881), and Cameroon (EPI_ISL_1790107, EPI_ISL_1790108, EPI_ISL_1790109). The most similar of these relatives is EPI_ISL_3316023, which was sampled on 11 January 2021 in California and represents the direct ancestor of the minor variant on the phylogeny. The only mutation separating these genomes is T28272A, which is a reversion away from an Epsilon-defining mutation. There are no sequenced closely related Epsilon genomes from NYC, although this variant was present at a prevalence around 1% in NYC during the first two weeks of 2021 in NYC ^22^.

### Four-gamete tests of recombination

A preliminary inquiry of the genome sequencing data from the S (12 contiguous read fragments) and nucleoprotein (3 contiguous read fragments) regions was suggestive of recombinant genome fragments within the named contact partner. To determine whether pairs of polymorphic sites within individual read fragments displayed evidence of recombination we employed three different four-gamete based recombination detection tests: PHI ^23^, MCL, and R^2^ vs Dist ^24^ (Table 2). The power of each of these tests to detect recombination was seriously constrained by the short lengths of the read fragments and the low numbers of both variant-defining sites and other polymorphic sites with minor allele frequencies >1% within each of the fragments. Only three of the 15 read fragments (read fragments 6 and 8 in the S-gene and read fragment 3 in the N-gene) encompassed two or more of the variant-defining sites that were expected to provide the best opportunities to detect recombination. Nevertheless, pairs of sites within four read fragments in the S gene (positions 23123–24467 covering fragments 7, 8, 9 and 10) and one read fragment in the nucleoprotein gene (positions 28986–29378 covering fragment 3) exhibited signals of significant phylogenetic incompatibility with at least two of the three tests (*p* < 0.05): signals which are consistent with recombination. The only read fragment for which evidence of recombination was supported by all three tests was fragment 3 in the N-gene: a fragment that was one among only three that contained multiple variant-defining sites. Eight of the fifteen analyzed read-fragment alignments exhibited no signals of recombination using any of the tests, which is unsurprising given the lack within these fragments of both variant-defining substitutions and polymorphic sites with minor allele frequencies greater than 1%.

**Table 2:** Four gamete recombination test results for 15 sets of aligned read fragments in the S (spike) and N (nucleoprotein) genes.

| Gene | Fragment | Start<sup>1</sup> | End<sup>1</sup> | Recombination test *p*-value |  |  |
| --- | --- | --- | --- | --- | --- | --- |
|  |  |  |  | PHI | MCL | R<sup>2</sup> vs Dist |
| S | 1 | 21354 | 21730 | >0.9 | >0.9 | >0.9 |
|  | 2 | 21658 | 22038 | >0.9 | >0.9 | >0.9 |
|  | 3 | 21962 | 22346 | >0.9 | >0.9 | >0.9 |
|  | 4 | 22263 | 22650 | 0.682 | 0.393 | 0.132 |
|  | 5 | 22517 | 22903 | >0.9 | 0.401 | 0.142 |
|  | 6 | 22798 | 23212 | <0.001 | 0.355 | 0.3 |
|  | 7 | 23123 | 23522 | >0.9 | <0.001 | 0.005 |
|  | 8 | 23444 | 23847 | <0.001 | 0.317 | 0.008 |
|  | 9 | 23790 | 24169 | >0.9 | 0.047 | 0.03 |
|  | 10 | 24079 | 24467 | >0.9 | 0.003 | <0.001 |
|  | 11 | 24392 | 24789 | <0.001 | 0.071 | 0.477 |
|  | 12 | 24697 | 25076 | 0.741 | 0.155 | 0.208 |
| N | 1 | 28395 | 28779 | 0.491 | 0.482 | 0.459 |
|  | 2 | 28678 | 29063 | >0.9 | 0.868 | 0.628 |
|  | 3 | 28986 | 29378 | <0.001 | <0.001 | <0.001 |

<sup>1</sup>Position relative to the Wuhan-Hu-1 reference strain

### Targeted sequencing for recombination detection

The four gamete tests on genomic sequencing data is limited by the short length of amplified fragments. To obtain data from longer sequence fragments, we PCR-amplified three regions of the genome from the original nucleic acid extracts, cloned them, and then sequenced individual clones. These longer genomic fragments provide greater resolution for detecting recombination, compared with the short fragments from deep sequencing analysis, because they include more differentiating sites spread out farther across the genome.

The longest cloned region spanned 947 nt within the S gene and contained 5 nt substitutions differentiating the major and minor strains plus a variable site in the major variant. Of the 104 clones sequenced within this region, 60 (57.7%) were major strain haplotypes, 13 (12.5%) were minor strain haplotypes, whereas the remaining 31 clones (29.8%) contained both major and minor strain mutations, consistent with recombination (Figure 3A). We observed 11 distinct combinations of major and minor strain mutations across these clones, with two distinct haplotypes present in 6 clones apiece. Most haplotypes are consistent with only a single recombination breakpoint, though we did observe clones consistent with 2 or 3 breakpoints.

**Figure 3.**
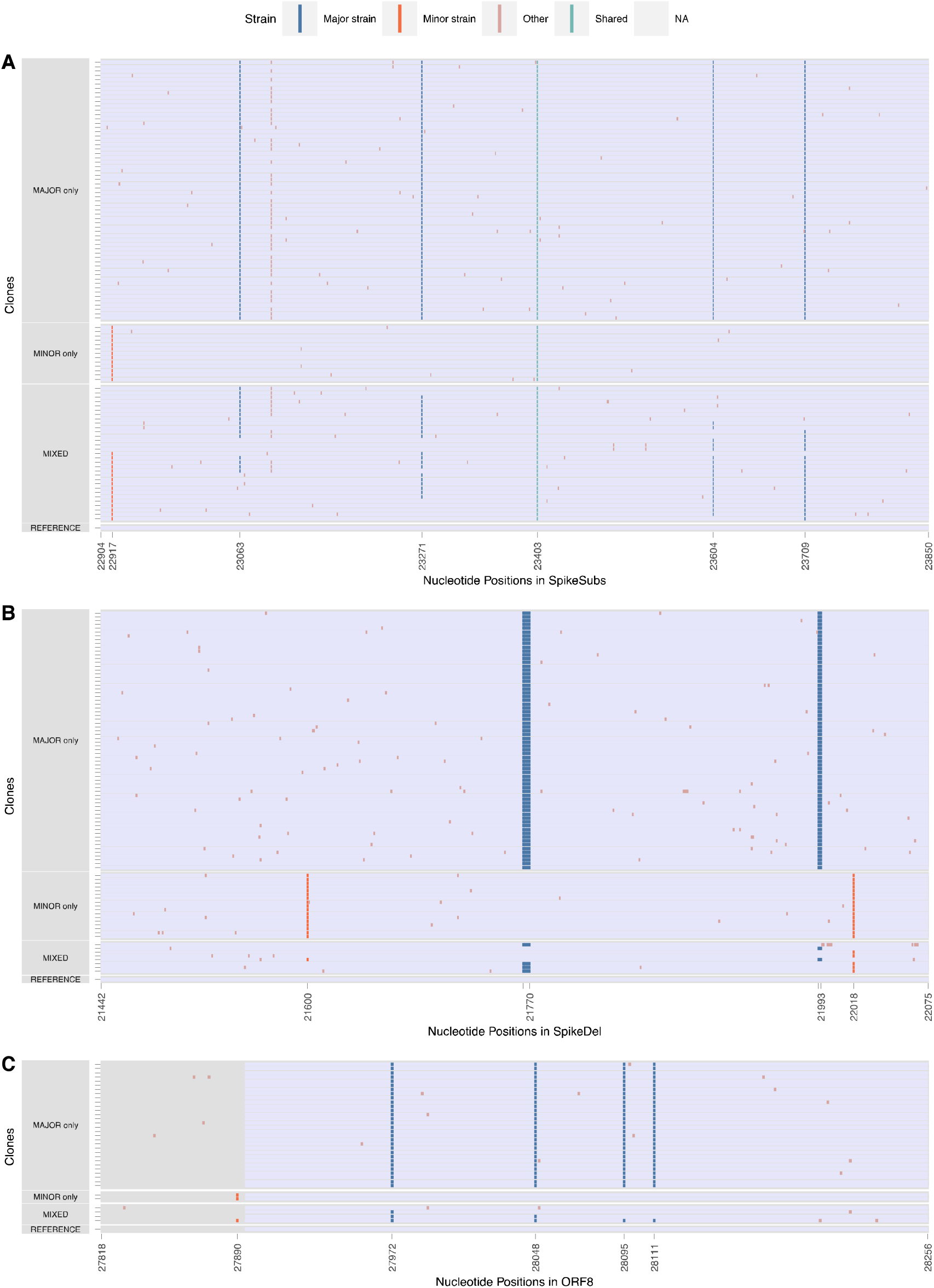
Major, minor, and mixed alleles in cloned sequences. Clones in S-substitution (S_Subs), S-Deletion (S_Del), and ORF8 were sequenced. Major and Minor allele positions are defined by the variant calling analysis performed on Galaxy. For each of the cloned region here, there are clones with only major alleles, minor alleles, and mix of both alleles. Out of the three cloned regions, the S-substitution clones has the highest frequency of mixed variants.

**Figure 4.**
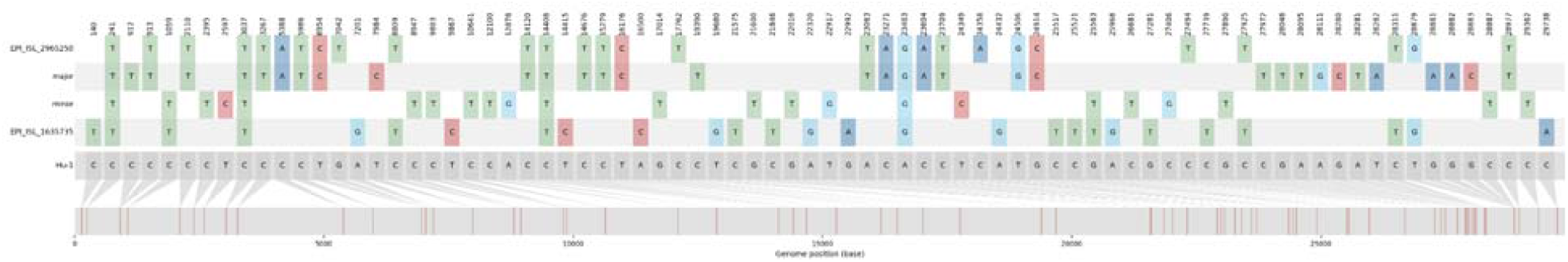
The nucleotide variation present in the major, minor, and putative recombinant strains. The distribution of the nucleotide variation found in the major, minor, B.1.526 (EPI_ISL_1635735), and single putative recombinant (EPI_ISL_2965250) strains relative to the reference genome (Wuhan Hu-1; bottom grey sequence).

The second cloned S region spanned 658 nt in S including the Δ69-70 and Δ144 deletions characteristic of the major strain and two 2 substitutions in the minor strain. Of the 93 clones sequenced, 69 (74.1%) were major strain haplotypes, 17 (18.3%) were minor strain haplotypes, and 7 (7.5%) were mixed haplotypes (Figure 3B). Five of these mixed haplotypes contained only one of the two deletions. Unlike in the primary sequencing analyses where the Δ69-70 and Δ144 deletions were present in >98% of sequences, Δ69-70 was observed in only 72 (77.4%) clones and Δ144 was observed in only 71 (76.3%). These frequencies are consistent with the frequency of the other major strain substitutions in the primary sequencing analysis.

The third, and shortest, cloned region spanned 476 nt of ORF8, surrounding 4 substitutions defining the major strain and 1 minor strain substitution. Of the 36 cloned sequences, 30 (83.3%) had the major strain haplotype, 2 (5.6%) had the minor variant haplotype, and 4 (11.1%) had mixed haplotypes consistent with recombination (Figure 3C). Notable, the discriminating substitutions only span 223 nt of this region.

### Consistency between cloning and genome sequencing analyses

*In-vitro* recombination can be introduced by reverse-transcription and PCR amplification, which are part of both genome sequencing and cloning protocols ^25^. These *in-vitro* effects have a strong stochastic component and would result in substantially different recombinant haplotype frequencies across different extracts and PCR experiments. To determine the extent to which these protocols could have led to biased inference of recombination, we compared the haplotype frequencies across the four extracts from NYCPHL-002461, which had each independently been subjected to reverse transcription and PCR amplification, and the frequency of these haplotypes in the cloning experiment, which included PCR amplification.

Within the S gene substitution region encompassing nucleotide positions 23604–23709, the major haplotype was present between 76.4% and 78.6%, and the minor haplotype was between 13.7% and 15.4% (Supplementary Table 1). The recombinant haplotype positions 23604A and 23709C was present at 3.9% allele frequency (standard deviation of 0.34% across extracts), whereas recombinant haplotype 23604C and 23709T was present at 4.3% (standard deviation of 0.37% across extracts). Although the haplotype frequencies among extracts were significantly different (*p=*0.029; chi-square test), the magnitude of these differences were unremarkable. Furthermore, there was no significant difference between the frequency of these haplotypes in cloning experiment and extracts (*p*=0.190 versus -A; *p*=0.189 versus -B; *p*=0.357 versus -C; *p*=0.206 versus -D; Fisher’s Exact Test).

A similar pattern was observed within the ORF8 region at nucleotide positions 27972–28111, which included four discrimination sites: 27972, 28048, 28095, and 28111 (Supplementary Table 2). The predominant recombinant haplotypes were consistent across the four extracts, and the frequencies differed only slightly (*p*=0.077; chi-square test). As in S, the frequency of these recombinant haplotypes in the cloning experiment was not significantly different from any of the extracts (*p*=0.405 versus -A; *p*=0.413 versus -B; *p*=0.199 versus -C; *p*=0.408 versus -D; Fisher’s exact test).

Hence, *in-vitro* recombination induced by either reverse-transcription or PCR amplification, does not appear to have been the dominant contributor to the recombinant haplotype distribution reported here.

### Search for transmission of a circulating recombinant

To determine whether there was onward transmission of a recombinant descendent of these major and minor strains, we queried the 27,806 genomes sequenced by NYC public health surveillance and deposited to GISAID through 05 September 2021. We tested these genomes for mosaicism (3SEQ; ^26^; with Dunn-Sidak correction for multiple comparisons) of the major and minor strains; however, we were unable to reject the null hypothesis of non-reticulate evolution for any of these genomes. There is no evidence of an Alpha/Epsilon recombinant that circulated in New York City.

Since the Dunn-Sidak correction done in the 3SEQ analysis applies a conservative type-1 error threshold of 0.05, we reran the analysis using a more permissive threshold of 0.25 (see methods) and were able to reject the null hypothesis for a single genome (EPI_ISL_2965250; *p*=2.24×10^−6^ and Dunn-Sidak corrected *p*=0.117). Although this genome contains many of the mutations characteristic of the Alpha variant throughout the genome, it does not possess mutations unique to the major strain nor any Epsilon-specific mutations. Rather, within the putative recombinant regions, the EPI_ISL_2965250 genome has C8809T, C27925T, C28311T, and T28879G. All of these mutations are characteristic of the B.1.526 Iota-variant, prevalent in NYC in early 2021. Therefore, this genome is likely not a descendant of the major and minor strains. Instead it appears to be a recombinant descendant of Alpha and Iota viruses.

## DISCUSSION

Here, we report evidence of intra-host recombination of SARS-CoV-2 within a single individual superinfected with Alpha and Epsilon viral variants during the second COVID-19 wave in New York City in early 2021. Because recombinant viruses can be successfully generated and transmitted^15^ between humans, this finding underscores their potential relevance to the future of the COVID-19 pandemic.

The presence of major and minor strains described within the superinfected individual are unlikely to be the result of bioinformatics error, contamination, or experimental artifacts. The degree of evolutionary divergence of each of the strains from other available SARS-CoV-2 genomes is consistent with viruses circulating at the time of their January 2021 sampling dates. Moreover, the major strain genome is identical to contemporaneously sampled genomes from both a named contact and strains circulating in the country from which they had both recently visited. No closely related genome to the minor strain was ever sequenced by NYC DOHMH, lessening the probability of a contaminated sample. Given the relatively low sequencing coverage in NYC in January 2021 and low prevalence of the Epsilon variant, around 1% in NYC at the time, it is not unexpected that a closely related genome would not be observed. Furthermore, the major and minor variants were both present in all four extractions of the two aliquots at similar frequencies, indicating that any contamination, if present, would need to have occurred in the original sample swab.

The timing of this superinfection is important, because January 2021 was the peak of the second COVID-19 wave in NYC, a time when numerous variants were circulating and immediately prior to the vaccination roll-out campaign. Hence, January 2021 in NYC represents not only the height of potential for superinfection risk, but also a location where its existence would be most apparent due to the co-circulation of numerous genetically distinct viral variants.

There remain unexplained patterns in the genome sequencing data from the superinfected individual. Evidence of a major and minor strain was not apparent at the S deletions Δ69/70 and Δ144 in the genome sequencing, but the cloning analysis showed major and minor alleles at these sites at the expected frequencies. Therefore, it is possible that the ARTIC protocol preferentially sequenced templates containing these deletions, giving a false impression of their predominance in the genomic analysis. Also of interest is the A28272T mutation in the minor strain, which is either a reversion or potential sequencing artifact. If the base-call at position 28272 in the minor variant is erroneous, then the minor strain would be identical to a virus sampled contemporaneously in California, where the Epsilon variant was first discovered and likely originated.

Laboratory induced recombination is a common artifact during reverse-transcription and PCR ^27,28^. However, recombination is a pervasive feature of natural coronavirus infection, as it has been observed in bats, camels, and humans ^2,15,29,30^. One would *a priori* expect to find recombinant viruses in a SARS-CoV-2 superinfected individual. Therefore, it is unlikely that the entirety of the signal for recombination reported here is due to reverse-transcription or PCR-induced recombination. A consistent signal for recombination was observed in the four whole genome sequencing analyses and in cloned-fragment analysis, all suggesting the same recombinant haplotypes present at high frequency.

Our search for Alpha/Epsilon variant recombinants in NYC did not identify genomes that would suggest onward transmission of either of the major or minor strains derived here, or a recombinant offspring. This lack of onward transmission is not surprising, given that the initial index case was contacted by NYC DOHMH personnel and their named contact (the superinfected case) received a prompt COVID-19 diagnosis and was advised to self-isolate.

It is likely that superinfection with SARS-CoV-2 is more common than has been described in the literature, especially given the documentation of circulating recombinant strains of the Alpha variant in the United Kingdom ^15^. Recombinant virus can only be produced within a superinfected individual. That said, we caution against assuming superinfection before potential issues of contamination, poor-quality sequencing, or bioinformatics errors have been appropriately dealt with.

The high number and genomic variability recombinant haplotypes that we have identified within a single superinfected individual suggests that recombination is perpetually occurring within SARS-CoV-2 infections. Whether recombination will play a role in the emergence of novel SARS-CoV-2 variants is an open question. Reduced incidence due to vaccine-induced and naturally-acquired immunity would lower the opportunity for superinfection, and the homogenizing effect of variant-driven selective sweeps (as seen in the Delta and Omicron variants ^31^) will lessen the potential for biological innovation in a recombinant genome. Nonetheless, SARS-CoV-2 molecular surveillance should actively monitor for the emergence of a recombinant variant.

## MATERIALS AND METHODS

### Extraction and sequencing

Nasopharyngeal specimens positive for SARS-CoV-2 with Ct < 32 were submitted to NYC PHL for sequence analysis by the NYC DOHMH through the COVID Express clinics. The NYCPHL-002130 and -002461 specimens had Ct values of 19 and 20 cycles, respectively, which allowed for sequencing at NYC PHL. Each specimen was split into separate extraction and archive aliquots. Nucleic acid extraction was performed using the KingFisher Flex Purification System (Thermo Fisher Scientific) from the extraction aliquot. Eleven µL of extract was used to anneal with random hexamers and dNTPs (New England Biolabs Inc., NEB) and reverse transcribed with SuperScript IV Reverse Transcriptase at 42 ºC for 50 min. The cDNA product was amplified in two separate multiplex PCRs with ARTIC V3 primer pools (Integrated DNA Technologies) in the presence of Q5 2x Hot Start Master Mix (NEB) at 98 ºC for 30 s, and 35 cycles of 98 ºC for 15 s and 65 ºC for 5 min. The two PCR products were combined and were purified with Agencourt Ampure XP magnetic beads (Beckman Coulter) at a 1:1 sample-to-bead ratio. The bead-cleaned PCR products were quantified using a Qubit 3.0 fluorometer (Thermo Fisher Scientific). Standard protocol was used for library preparation in the NEBNext Ultra II Library Preparation workflow using 90 ng of PCR product (NEB). In short, the ARTIC PCR products were used in an end-repair reaction, which added a 5’-phosphate group and a dA-tail, in a reaction for 30 min at 20 ºC. The reaction was heat inactivated for 30 min at 65 ºC. NEBNext Adaptor was ligated at 25 ºC for 30 min and cleaved by USER Enzyme at 37 ºC for 15 min. The product was Agencourt Ampure XP bead-purified at a ratio of 0.6x sample:beads. The bead-cleaned, end-ligated amplicons were subjected to a 6-cycle PCR reaction with NEBNext Ultra II Q5 Master Mix in the presence of NEBNext Multiplex Oligos for Illumina (NEB), which added a sample-specific 8-base index and Illumina P5 and P7 adapters for sequencing on Illumina instruments. The product was purified with Ampure XP beads at a 0.6x sample:bead ratio and quantified, normalized and pooled at equimolar concentration with other libraries, followed by loading onto the Illumina MiSeq sequencing instrument using V3 600-cycle reagent kit, with a V3 flow cell for 250-cycle paired-end sequencing (Illumina). For NYCPHL-002461, the same “extraction” specimen aliquot was used for a second extraction, Extract B. Extracts C and D were independent extractions, but from the “archived” specimen aliquot. As such, the first extract (A), and extracts B, C, and D were independent samples which underwent independent reverse transcription, ARTIC PCR, library preparation, and sequencing reactions.

Potential *in vitro* recombination that occurred during the four independent extractions, reverse-transcription reactions, and library preparation procedures, such as PCR amplification, would require the events to occur independently at the same stage four times in order to produce the same proportions of Major and Minor variant haplotypes in the high-throughput sequencing data. To account for *in vitro* recombination, regions where long complete reads span across major and minor variants in close proximity, <105 nucleotide bases, were examined across all genome alignments of the four NYCPHL-002461 replicates. Reads with SAM (Sequence Alignment Map) Flags 81,83,97,113,145,147,161,177,2129 are included in the analysis. Reads with other flags are excluded from the analysis or are not found in the alignment files. Additionally, reads without a combination of major or minor alleles are excluded. All unique haplotypes at major and minor variant positions are grouped together. Relative frequencies of the haplotypes are calculated for each region of all four extractions.

### Cloning

Three regions of the SARS-CoV-2 genome from NYCPHL-002461 were cloned. Two contained non-overlapping regions of the S gene and were designated S_Subs (positions 22882–23873) and S_Del (positions 21421–22098). The third region included part of the ORF8 gene (positions 27798–28280).

To perform the annealing step for reverse transcription, 3 µl of the NYCPHL-002461 nucleic acid extract was combined with reverse primer and dNTPs at final concentrations of 154 nM and 769 µM, respectively. Annealing was performed by heating to 65°C for 5 minutes then cooling to 4°C. The reverse primer sequences used for the reverse transcription are as follows: S_Subs primer-R is 5□-CTATTCCAGTTAAAGCACGGTTT, S_Del is 5□-AGGTCCATAAGAAAAGGCTGAGA and ORF8 is 5□-GAGACATTTAGTTTGTTCGTTTA. An elongation mix containing 200 units of SuperScript IV Reverse Transcriptase (Invitrogen) and 40 units of RNaseOUT Recombinant Ribonuclease Inhibitor (Invitrogen) was then added along with dNTPs at a final concentration of 200 µM. The resulting solution was heated to 55°C for 10 minutes then 80°C for 10 minutes.

Each cDNA target was PCR amplified using Platinum II Taq polymerase and its accompanying buffer (Invitrogen) supplemented with a final concentration of 200 µM dNTPs and 600 nM of each primer. The reverse primer sequences used for reverse transcription were included along with their corresponding forward primers: S_Subs primer-F is 5□-TCTTGATTCTAAGGTTGGTGGT, S_del is 5□-AGGGGTACTGCTGTTATGTCT and ORF8 is 5□-GCCTTTCTGCTATTCCTTGT. PCR was performed with the following cycling conditions: an initial hold at 94°C for 2 minutes, followed by 35 cycles at 94°C for 15 seconds, 60°C for 15 seconds and 68°C for 15 seconds. Cycling was followed by a final extension step at 72°C for 7 minutes. The PCR products were run on 2% agarose gels, excised and gel purified with the GenElute Gel Extraction Kit (Sigma). The gel-purified PCR products were cloned using the TOPO TA Cloning Kit for Sequencing (Invitrogen).

Individual colonies resulting from the transformation into chemically competent One Shot TOP10 *Escherichia coli* (Invitrogen) were picked and patch plated for sequencing. Rolling circle amplification and Sanger sequencing of the clones were performed by GeneWiz (New Jersey). All clones were sequenced with the M13 reverse primer. Due to its larger size, S_Sub clones were also sequenced with the M13 forward primer.

### Major and minor variant calling

We used the Galaxy SARS-CoV-2 variant calling pipeline for paired-end Illumina ARTIC amplicon data ^21^. Briefly, the workflow performs quality control, masks primer sites, maps reads to reference using *BMA-mem*, calls variants using *lofreq*, annotates them using *SNPEff*, and outputs tabular variant call files, thresholded on minimum allele frequency of 0.05. These variants are further visualized in a custom ObservableHQ notebook (https://observablehq.com/@spond/nyc-superinfection).

### Alignment and phylogenetic inference

SARS-CoV-2 genomes were downloaded from GISAID on 24 March 2021. Genomes assigned to the B.1.1.7 or B.1.429 Pangolin lineage ^32^ were aligned to the Wuhan-Hu-1 reference genome using the --6merpair option in Mafft v7.464 ^33^. We further refined these alignments to only those genomes sharing specific synapomorphies with the major and minor allelic variants from NYCPHL-002461: C241T, C3037T, C14408T, and A23403G for B.1.1.7 viruses and C8947T, C12110T, and C10641T for B.1.429 viruses. Separate maximum likelihood phylogenetic trees for B.1.1.7 (n=1176) and B.1.429 (n=807) were inferred in IQTree2 under a GTR+F+I model, with the additional NNI search option and a minimum branch length of 1e-9 substitutions/site^34^.

### Molecular clock inference

To determine whether the major and minor allelic variants were contemporaneous with the date of sampling, we estimated clock trees for the B.1.1.7 and B.1.429 phylogenies in TreeTime v0.8.0^35^. We fixed the clock rate to 8×10^−4^ substitutions/site/year under a skyline coalescent model (per NextStrain default parameters for SARS-CoV-2). These trees were also used to estimate the time to most recent common ancestor of the allelic variants and their closest relatives.

### Four-gamete tests for recombination

When sequences evolve in the absence of both recombination and convergent mutations it is expected that, for any pair of polymorphic sites where one of the sites has either nucleotide X or Y and the other has either nucleotide A or B, no more than three of the four possible combinations of nucleotides (or gametes) at the two sites (i.e. XA, XB, YA and YB) should ever be observed. Given that in reality convergent mutations are always possible, four gamete tests of recombination attempt to detect situations where the numbers of site pairs where all four combinations of nucleotides are observed exceed that expected due to convergent mutations in the absence of recombination. We tested 15 multiple sequence alignments, each containing all observed unique read fragment sequences spanning the S-gene (12 fragments) and N-gene (3 fragments) with three different four gamete tests: (1) the PHI test (implemented in RDP5 ^23,36^) which considers sites with more than two alternative nucleotide states and uses a permutation-based test to determine whether detected site pairs displaying all four gametes display a degree of spatial clustering along the sequence that is significantly higher than would be expected in the absence of recombination; (2) the MCL recombination detection test (implemented in the pairwise component of the LDHat package; ^24^) which uses an approximate maximum likelihood method to infer the population scaled recombination rate needed to explain the observed numbers of site pairs with four gametes and then tests for significant deviation of the inferred recombination rate from zero using a permutation test, and (3) the RvsDist test (implemented in pairwise component of LDHat) which determines the correlation between the R^2^ measure of linkage disequilibrium between site pairs with four gametes with the physical distance in nucleotides between the site pairs ^24^ and uses a permutation test to detect significant deviations from the expected degree of correlation in the absence of recombination. For both the MCL and RvsDist tests we used a minor allele frequency cutoff of 0.01.

### Population level recombination detection

We downloaded all SARS-CoV-2 sequences from GISAID that were deposited by 5 September 2021 ^37^ and analyzed the 27,806 genomes sequenced by the NYC PHL and Pandemic Response Lab (PRL) from specimens collected from NYC residents. These genomes were aligned to the Wuhan-Hu-1 reference genome (Genbank accession NC_045512.2) using MAFFT v7.453 (options --auto --keeplength -- addfragments) ^33^.

To determine whether there was any onward transmission of a major-minor strain recombinant, we used 3SEQ v.1.7 ^26^ as a statistical test for recombination in the NYC data. 3SEQ interrogates triplets of sequences for signals of mosaicism in a sequence given two ‘parental’ sequences. We interrogated each of the 27,806 NYC PHL and PRL-generated genomes for mosaicism given the major and minor strains as parents. The resulting *p*-values are Dunn-Sidak corrected for multiple comparisons (n=55612), and we tested for mosaicism at *p*-value thresholds 0.05 and 0.25. The single nucleotide differences between a putative recombinant and the major and minor strains were visualized using snipit (https://github.com/aineniamh/snipit).

## Data availability

The data analyzed as part of this project were obtained from the GISAID database and through a Data Use Agreement between NYC DOHMH and the University of California San Diego. We gratefully acknowledge the authors from the originating laboratories and the submitting laboratories, who generated and shared via GISAID the viral genomic sequence data on which this research is based. A complete list acknowledging the authors who submitted the data analyzed in this study can be found in Data S1.

Trimmed, host-depleted viral sequencing data and cloned sequence fragments have been submitted to NCBI (accession numbers pending).

## Data Availability

The data analyzed as part of this project were obtained from the GISAID database and through a Data Use Agreement between NYC DOHMH and the University of California San Diego. We gratefully acknowledge the authors from the originating laboratories and the submitting laboratories, who generated and shared via GISAID the viral genomic sequence data on which this research is based. A complete list acknowledging the authors who submitted the data analyzed in this study can be found in Data S1.
Trimmed, host-depleted viral sequencing data and cloned sequence fragments have been submitted to NCBI (accession numbers pending).

## Acknowledgements

J.O.W. acknowledges funding from the National Institutes of Health (AI135992 and AI136056). D.P.M was funded by the Wellcome Trust (222574/Z/21/Z). PKQ is funded by a Crick African Network Fellowship. T.I.V. is funded by a Branco Weiss Fellowship. S.L.K.P and A.N were supported in part by a grant from the National Institutes of Health (AI134384). This work was supported (in part) by the Epidemiology and Laboratory Capacity (ELC) for Infectious Diseases Cooperative Agreement (Grant Number: ELC DETECT (6NU50CK000517-01-07) funded by the Centers for Disease Control and Prevention (CDC). Its contents are solely the responsibility of the authors and do not necessarily represent the official views of CDC or the Department of Health and Human Services.

## SUPPLEMENTAL TABLES

**Supplementary Table 1.** Recombinant haplotype frequencies across separate extractions, reverse transcription, and PCR amplification in high-throughput sequencing for S-Substitution (S_Sub) region, nucleotide positions 23604 and 23709.

| Haplotypes |  |  | Count (Frequencies) |  |  |  |
| --- | --- | --- | --- | --- | --- | --- |
| Type | 23604 | 23709 | NYCPHL-002461-A | NYCPHL-002461-B | NYCPHL-002461-C | NYCPHL-002461-D |
| MAJOR | A | T | 3633 (78.57%) | 4095 (76.43%) | 3778 (76.40%) | 4745 (78.24%) |
| MINOR | C | C | 634 (13.71%) | 776 (14.48%) | 762 (15.41%) | 851 (14.03%) |
| MIX | A | C | 162 (3.50%) | 234 (4.37%) | 191 (3.86%) | 237 (3.91%) |
| MIX | C | T | 195 (4.22%) | 253 (4.72%) | 214 (4.33%) | 232 (3.83%) |
| Depth |  |  | 4624 (100%) | 5358 (100%) | 4945 (100%) | 6065 (100%) |

**Supplementary Table 2.** Recombinant haplotype frequencies across separate extractions, reverse transcription, and PCR amplification in high-throughput sequencing for ORF8 region, nucleotide positions 27972, 28048, 28095, 28111.

| Positions |  |  |  |  | Count (Frequencies) |  |  |  |
| --- | --- | --- | --- | --- | --- | --- | --- | --- |
| Haplotype | 27972 | 28048 | 28095 | 28111 | NYCPHL-002461-A | NYCPHL-002461-B | NYCPHL-002461-C | NYCPHL-002461-D |
| MAJOR | T | T | T | G | 1382 (70.22%) | 1164 (69.83%) | 1110 (68.90%) | 1873 (70.26%) |
| MINOR | C | G | A | A | 381 (19.36%) | 305 (18.30%) | 347 (21.54%) | 505 (18.94%) |
| MIX | T | G | A | A | 57 (2.90%) | 40 (2.40%) | 31 (1.92%) | 77 (2.89%) |
| MIX | C | T | T | G | 50 (2.54%) | 60 (3.60%) | 55 (3.41%) | 66 (2.48%) |
| MIX | T | T | A | A | 40 (2.03%) | 43 (2.58%) | 28 (1.74%) | 48 (1.80%) |
| MIX | C | G | T | G | 34 (1.73%) | 28 (1.68%) | 18 (1.12%) | 45 (1.69%) |
| MIX | T | T | T | A | 8 (0.41%) | 7 (0.42%) | 6 (0.37%) | 12 (0.45%) |
| MIX | C | G | A | G | 4 (0.20%) | 6 (0.36%) | 4 (0.25%) | 12 (0.45%) |
| MIX | T | G | T | G | 6 (0.30%) | 8 (0.48%) | 7 (0.43%) | 15 (0.56%) |
| MIX | C | T | A | A | 1 (0.05%) | 3 (0.18%) | 3 (0.19%) | 5 (0.19%) |
| MIX | C | G | T | A | 1 (0.05%) | 1 (0.06%) | NULL | NULL |
| MIX | T | T | A | G | 1 (0.05%) | 1 (0.06%) | 2 (0.12%) | 7 (0.26%) |
| MIX | C | T | A | G | 1 (0.05%) | 1 (0.06%) | NULL | NULL |
| MIX | T | G | T | A | 1 (0.05%) | NULL | NULL | NULL |
| MIX | C | T | T | A | 1 (0.05%) | NULL | NULL | 1 (0.04%) |
| Depth |  |  |  |  | 1968 (100%) | 1667 (100%) | 1611 (100%) | 2666 (100%) |

